# Diagnosis, course and outcomes of comorbidity of tuberculosis, opportunistic respiratory infections and COVID-19 in patients with advanced HIV infection with immunodeficiency

**DOI:** 10.1101/2025.04.11.25325673

**Authors:** A.V. Mishina, V.Yu. Mishin, I.A. Vasilieva, I.V. Shashenkov

**Affiliations:** FGBOU VO “Russian University of Medicine” of the Ministry of Health of Russia, Moscow, Russia; FGBU “National Medical Research Center for Phthisiopulmonology and Infectious Diseases” of the Ministry of Health of Russia; GBUZ “Prof. G.A. Zakharyin Tuberculosis Clinical Hospital No. 3” of Moscow Department of Health, Moscow, Russia

**Keywords:** comorbidity, tuberculosis, opportunistic respiratory infections, COVID-19, HIV infection, immunodeficiency, microbiological diagnostics, molecular genetic diagnostics, immunological diagnostics, radiation diagnostics

## Abstract

**Purpose:** To study the features of the diagnosis, course and outcomes of comorbidity of tuberculosis (TB), opportunistic respiratory infections (ORI) and COVID-19 in patients with advanced HIV infection with immunodeficiency (ID).

**Materials and methods:** The prospective two-year follow-up study included 58 patients aged 26-56 years who were randomized into 2 groups. The 1st group included 29 patients with TB, ORI and COVID-19 comorbidity, of 4B stage of HIV infection in the progression phase and in the absence of ART. Group 2 - 29 patients selected according to the “copy-pair” principle, identical to patients of Group 1, but without COVID-19.

**Results:** The comorbidity of TB, ORI, and COVID-19 in patients with advanced HIV infection is characterized by marked immunodeficiency and generalization of tuberculosis with multiple extrapulmonary manifestations and high levels (more than 70%) of MDR and BDR. Significant improvement in patients of the 1st and 2nd groups was established in 24.1% and 20.7% of cases, progression in 31.0% and 41.4%, respectively, death in 41.3% and 37.9%. Progression and death were associated with lack of treatment adherence, drug dependence, severe generalized TB and ORI progression.

**Conclusion:** Patients with TB, ORI and COVID comorbidity in the late stages of HIV infection with ID represent a high risk group for severe disease and death, due to, among other things, social maladjustment and lack of adherence to examination and treatment, active examination of such patients. It is required to establish an etiological diagnosis necessary for their emergency hospitalization in a specialized infectious diseases hospital for adequate comprehensive treatment.

## Introduction

The comorbidity of tuberculosis (TB) and HIV infection is a classic model of the simultaneous development of several pathologies, where, during development of immunodeficiency (ID) opportunistic respiratory infections (ORI) join, is accompanied by high mortality.

Recently, a number of works have been published covering the features of the clinic, diagnosis and treatment in patients with comorbidity of HIV infection with COVID-19 [1-5] and TB with COVID-19 [6-8], that present data on the features of clinical and radiological manifestations of comorbid pathology and the effect of COVID-19 on clinical manifestations and course of HIV infection or TB. The comorbidity of TB, ORI and COVID-19 presents certain problems in patients in the later stages of HIV infection with ID in terms of diagnosis, assessment of clinical and radiological manifestations, peculiarities of the course, timely adequate etiological treatment and outcomes, while there are relatively few works devoted to that problem [9-11]

### Purpose

To study the features of diagnosis, course and outcomes of TB, ORI and COVID-19 comorbidity in patients with advanced HIV infection with ID.

## Materials and methods

The materials include data of a prospective cohort study and a dynamic two-year follow-up study of 58 patients which were divided into the 1st (main) and the 2nd group (for comparison). The 1st group consisted of 29 patients with TB comorbidity with isolation of Mycobacterium tuberculosis (MBT), ORI and COVID-19, 4V stage of HIV infection in progression phase and in absence of ART at age of 26-56 years, number of men were 19 (65.5±8.8%) and women were 10 (34.5±8.8%). The 2nd group included 29 similar patients, selected on the basis of the “copy-pair” principle and completely identical to patients of the 1st group with practically similar age, gender, social and clinical-laboratory parameters, but without COVID-19.

Culture of MBT from patients was obtained from diagnostic material of the respiratory tract (sputum, bronchoalveolar lavage, biopsy material obtained from bronchoscopy and punctures of the intrathoracic lymph nodes) and other organs (blood, urine, feces and peripheral lymph node punctures) when sowing on a dense Lowenstein-Jensen medium and in the BACTEC MGIT 960 automated system and with determination of drug resistance of the obtained culture to anti-tuberculosis drugs (ATD) by the method of absolute concentrations [24, 25]. For the diagnosis of pathogens of ORI and the diseases provoked by them, such as lung mycobacteriosis (*Mycobacterium nontuberculosis)*, bacterial pneumonia (*Streptococcus pneumoniae (S. pneumoniae), Haemophilus ifluenzae (H. ifluenzae), Staphylococcus aureus (S. aureus))*, atypical ppneumonia (*Legionella pneumophila (L. pneumophila), Mycoplasma pneumoniae (M. pneumoniae)* и *Chlamydophila pneumoniae (C. pneumoniae))*, candidal pneumonia (*Candida albicans (C. albicans))* and viral pneumonia (*Herpesvirus Simplex* type 1 or *Cytomegalovirus Humanis)*, bacteriological, virological, immunological methods and polymerase chain reaction (PCR) diagnostic material [26] were applied. For evaluation of SARS-CoV-2, the RNA amplification with reverse transcription and fluorescence detection by real-time polymerase chain reaction (PCR) of the material was applied for the etiological diagnosis of COVID-19. All patients underwent comprehensive clinical, laboratory, immunological (determination of CD4+ lymphocytes number by flow cytofluorimetry and viral load using number of copies of HIV RNA in the peripheral blood) and radiation studies, including computed tomography of the chest organs (CT CO), magnetic resonance tomography (MRI) and ultrasound (US) of the internal organs Statistical data processing was carried out using the Microsoft Office Excel 2019 program with calculation of the group mean value and the standard error of the mean, confidence interval (CI). The significance test p was determined from the Student’s table. Differences between the arithmetic means were considered significant at p <0.05.

## Results

In all 58 patients of group 1 and group 2, HIV infection was the first disease and at the time of diagnosis 42 (72.4±5.9%) patients had recognized parenteral route of infection and 16 (27.6±5.9%) had sexual infection route. All of them were registered with an AIDS center, which was practically not visited due to social maladjustment and lack of adherence to examination and treatment and practically did not take ART. All patients were addicted to drugs, consumed alcoholic beverages and smoked tobacco products. All patients were diagnosed with concomitant diseases: viral hepatitis B or C and chronic obstructive pulmonary disease (COPD) - in 39 (67.2±6.2%).

In the observed groups, the number of CD4+ lymphocytes in 1 μL of blood practically did not differ from each other. In the 1st group, the CD4+ lymphocyte count was in the range of 50-30 cells/μL of blood was in 10.3±5.6% of patients, 29-20 - in 31.0±8.5%, 19-10 - in 34.5±8.8% and less than 9 - in 24.1±7.9%, while in the 2nd group, respectively: in 13.8±6.4%, 34.5±8.8%, 31.0±8.5% and 20.7±7.5% (p> 0.05). The mean number of CD4+ lymphocytes in 1 μL of blood was also the same, in patients of the 1st group was 24.1±0.64 cells/μL of blood, and in the 2nd group - 29.7±0.54 (p> 0.05). At the same time, the viral load in patients in both groups observed was more than 500 000 RNA of HIV copies/mL of blood.

The frequency of various OLIs and the frequency of their pathogens in patients in the observed groups did not differ significantly from each other. Bacterial pneumonia caused by *S. pneumoniae* was diagnosed in group 1 in 34.5±8.8% of patients and in group 2 - in 27.6±8.2% caused by *H. influenzae*, respectively: in 24.1±7.9% and in 20.6±7.5% and caused by *S. aureus*, respectively: - in 13.8±6.4% and in 17.2±8.5% (p> 0.05) and SARS caused by *L. pneumophil*a, respectively: in 20.6±7.5% and in 17.2±8.5% (p> 0.05) *caused by M. pneumoni*ae, respectively: in 17.2±8.5% and in 13.8±6.4% (p> 0.05) *caused by C. pneumon*iae, respectively: 13.8±6.4% and 17.2±8.5% (p> 0.05). Pneumocystis pneumonia *caused by P. jiro*veci was diagnosed in the 1st group in 24.1±7.9% of patients, and in the 2nd - in 20.6±7.5% (p> 0.05), candidal pneumonia, respectively: in 31.0±8.5% and 34.5±9.0% (p> 0.05), viral pneumonia *caused by Herpes virus simplex* of type 1, respectively: in 26.7±8.3% and 24.1±7.9±7.9% (p> *0*.*05)* and by *Cytomegalovirus hominis*, respectively: in 20.6±7.5% and 17.2±8.5% (p> 0.05) and mycobacteriosis of the lungs *caused by M. aviu*mcomplex, respectively, in 31.0±8.5% and 27.6±8.2% (p> 0.05) of patients. At the same time, the combination of two OLIs was in 12 patients of the 1st group and 11 - the 2nd, and the combination of the three, respectively: in 3 and 4.

The clinical picture of the disease in patients of the 1st and 2nd group practically did not differ from each other and was characterized by a pronounced syndrome of intoxication and general inflammatory changes, as well as shortness of breath, cough, secretion of mucopurulent sputum and presence of diverse wheezing in the lungs, and in some cases, skin rashes, anosmia, dysgeusia and neurosensory hearing loss, hypoxemia, DIC syndrome, thrombosis and thromboembolism, and in some cases - Guillain-Barré syndrome.

On CT of CO in patients in the observed groups, a complex of a simultaneous combination of four pathological syndromes is visualized: dissemination, pleural pathology, increased pulmonary pattern and adenopathy. At the same time, the area of lung damage in patients of the 1st and 2nd groups was 80-100% and was almost comparable between the groups. It was not possible to differentiate these changes on CT of CO by specific pathologies in view of similarity of the CT signs, while diagnosis is possible to be established only with microbiological, virological and molecular genetic determination of the pathogens etiology of tuberculosis, ORI, COVID-19 and HIV infection.

In the observed groups of MBT sensitive to all ATD and monoresistant in patients were not detected, and multiresistant - in the 1st group - in 27.6±8.3% and in the 2nd - in 20.7±7.5% (p> 0.05) of patients. Most patients with MBT had multiple and broad drug resistance (MDR and BDR), respectively: 44.8±8.2% and 51.7±9.3% and 27.6±8.3% and 27.6±8.3% (p> 0.05) of cases.

The outcomes of comorbidity in patients in the observed groups practically did not differ from each other. So after 2 years of observation, no clinical cure was established. Significant improvement was achieved only with the use of ART and adequate chemotherapy of TB and ORI and was characterized by a decrease in immunodeficiency and viral load, as well as an improvement in clinical condition, a decrease in intoxication, inflammatory changes in the respiratory system and partial resorption of pathological changes in the lungs, which was established in patients of the 1st group in 24.1±7.9% of cases and the 2nd - in 20.77.5% (p> 0.05). Progression was established in group 1 in 31.0±8.% of patients and in the 2nd - in 41.4±9.1% (p□> 0.05) and death, respectively: in 41.3±9.1% and 37.9±9.0% (p□> *0*.*05*). Progression and death are associated with lack of adherence to treatment, drug dependence, severe and generalized TB, and ORI progression.

## Conclusion

Patients with TB, ORI and COVID comorbidity in the advanced stages of HIV infection with ID represent a high-risk group of severe disease and death not only due to infectious comorbidity, but also due to social maladaptation, as well as lack of adherence to examination and treatment. This necessitates the mandatory active examination of patients with TB and HIV infection, especially in its late stages and in the absence of ART, to establish a correct etiological diagnosis and with the aim of their emergency hospitalization in a specialized infectious hospital for timely complex treatment and reduction of mortality.

## Data Availability

All data produced in the present work are contained in the manuscript.

## Ethical approval

The study was approved by the local ethics committee of the Yevdokimov Moscow State University of Medicine and Dentistry (former name of the Russian University of Medicine, protocol №677, 7.10.2022). The approval and procedure for the protocol were obtained in accordance with the principles of the Helsinki Convention.

## Consent for publication

Written consent was obtained from the patients for publication of relevant medical information and all of accompanying images within the manuscript.

## Author Credentials

**Mishina Anastasia Vladimirovna** – Ph.D., Associate Professor of VAK, Acting Head of Department of Phthisiology and Pulmonology of N.A. Semashko Institute of Clinical Medicine, Assistant Professor of Department of Radiation Diagnostics of Institute of Dentistry of the FGBOU VO “Russian University of Medicine” of the Ministry of Health of Russia, Associate Professor - Consultant of GBUZ “Prof. G.A. Zakharyin TKB No. 3”;

**Mishin Vladimir Yuryevich** - Dr. Med. Sciences, Professor, Chief Scientist of. at FGBU “National Medical Research Center for Phthisiopulmonology and Infectious Diseases” of the Ministry of Health of Russia, Honored Scientist of the Russian Federation, Honored Doctor of the Russian Federation, Academician at AEN.

**Vasilyeva Irina Anatolyevna** - Dr. Med. Sciences, Professor, Director of the FGBU “National Medical Research Center for Phthisiopulmonology and Infectious Diseases” of the Ministry of Health of Russia;

**Shashenkov Ivan Vasilievich** - Assistant Professor at Phthisiology and Pulmonology Department of FGBU VO “Russian University of Medicine” of the Ministry of Health of Russia; Corresponding author.

